# Alterations in secondary structure and binding affinity from mutations in the interleukin-1 receptor antagonist

**DOI:** 10.1101/2024.07.13.24310367

**Authors:** Joshua Pillai

**Affiliations:** School of Biological Sciences, University of California, San Diego, 9301 S Scholars Dr, La Jolla, 92093, CA, USA; Department of Neurosciences, University of California, San Diego, School of Medicine, 9500 Gilman Dr, La Jolla, 92093, CA, USA

**Keywords:** DIRA, Mutations, IL1RN, IL-1Ra, Autoinflammatory Disease, Deficiency of Interleukin-1 Receptor Antagonist

## Abstract

Deficiency of Interleukin-1 receptor antagonist (DIRA) is a rare autosomal recessive autoinflammatory disorder that occurs during the neonatal period, manifesting a spectrum of symptoms including but not limited to osteomyelitis and pustulosis. This disease results from loss-of-function mutations in the *IL1RN* gene, encoding the IL-1 receptor antagonist (IL-1Ra). The dysfunction of IL-1Ra may arise from two mechanisms: (i) disruptions in the folding of its secondary structure, and (ii) impaired binding to its receptor, IL-1 receptor type 1. However, limited information exists detailing broad alterations in structure and binding affinity of known IL-1Ra variants. Herein, we used a validated protein prediction model to visualize 15 unique variants, and subsequently performed a structural analysis to elucidate the mutational impacts on secondary structure and binding affinity. Our analyses revealed reduced affinity at the 5 critical binding sites (W16, Q20, Y34, Q36, and Y147) along with missing secondary structures (β-sheets, α-helices) among the variants.

**Highlights:**

- Structural analyses revealed reduced binding affinity resulting from impacts in critical binding sites of IL-1Ra.
- This is the first study broadly evaluating the mutational impacts on structure and binding affinity of IL-1Ra.
- A validated *in silico* protocol was used to predict structural alterations of the IL-1Ra variants.

## 1. Introduction

Interleukin-1 receptor antagonist (IL-1Ra) is a critical member of the IL-1 family, having an important role in binding to IL-1 receptors [1]. IL-1Ra is secreted from numerous cell types, such as immune, epithelial, and stromal cells, and is known to modulate immune and inflammatory responses [2]. Specifically, IL-1Ra is known to competitively inhibit binding of IL-1α and IL-1β from receptors without having any detectable intracellular responses [3]. This protein is encoded by the *IL1RN* gene, spanning approximately 400-kb on chromosome 2, with 4 isoforms variants being previously reported [4]. Naturally, given its role in regulating inflammatory responses, IL-1Ra is important to have in balance with IL-1 as it relates to maintaining normal physiology of various tissues and organs [5]. If imbalanced, with an overproduction of IL-1 and/or underproduction of IL-1Ra, can lead to the development of disease, as reported for gastric cancer, rheumatoid arthritis, and the deficiency of Interleukin-1 receptor antagonist (DIRA) [5–6].

DIRA is a rare autosomal recessive disease present near or at birth, and is characterized by symptoms including but not limited to osteomyelitis, pustulosis, and systemic inflammation [7]. With a mortality rate of 25%, there are no known patients who have reached adulthood with DIRA without prior treatment [7]. The primary cause of this autoinflammatory disease is from loss-of-function mutations occurring in the *IL1RN* gene, leading to lower secretion of IL-1Ra. Ultimately, this results in the uncontrolled pro-inflammatory signaling of cytokines IL-1α and IL-1β, and eventually systemic inflammation in patients.

Within current literature, there have been 34 patients that have been reported up to date. Only one clinical trial (ClinicalTrials.gov ID: NCT00059748) has been reported including 9 patients from Aksentijevich et al., and the remaining in individual case reports [8]. Due to the varying levels of reporting of patients in literature and the small population size, it is challenging to model this disease clinically [9]. One major gap in literature is that there are no studies that have broadly investigated the known cytogenetic abnormalities and their pathogenic impacts on the resulting IL-1Ra proteins, which are critical in understanding the pathophysiology of DIRA. Ultimately, because of the clinical needs and current gap in literature for this life-threatening condition, we were motivated to thoroughly analyze all of its known genetic variants in the context of their resulting protein secondary structures and binding affinities.

We first began by screening prior literature and public sources to acquire the precise protein sequences from known genetic variants. Then, we conducted a comprehensive structural analysis of all variants using an *in silico* protein prediction model, identifying changes in the secondary structure and critical binding sites of the resulting cytokine. We validate these results by discussing the *in vitro* experiments that were previously performed from the studies reporting these variants. Lastly, we review the results and discuss future directions for this rare disease.

## 2. Materials and Methods

### 2.1. Protein Sequence Acquisition

All cytogenetic abnormalities of known DIRA patients have been displayed in Table 1. As of August 8, 2024, there have been 34 DIRA patients and 16 variants that have been reported within literature from 22 studies [8–29]. Of all genetic variants, we identified 6 unique nonsense, 3 frameshift, 3 large deletions, 1 splice-site, 1 in-frame deletion, and 2 compound mutations displayed in Table 1. We precisely obtained these counts from a manual search on the PubMed database with the key-words on August 8, 2024: “Deficiency of Interleukin-1 Receptor Antagonist” OR “DIRA”. After review of literature, we found that most protein sequences were not directly listed from the publications or their supplementary materials. However, all publications included information on alterations from the wild type (WT) reference mRNA sequence provided by the National Center for Biotechnology Information (NCBI). All variants were directly referred to from transcript variant 1 (NM_173842.3) (*n* = 15) and variant 2 (NM_173841.2) (*n* = 1) of the *IL1RN* mRNA. After manually creating the mutations in each WT sequence, we verified most sequences with the Infevers database (http://fmf.igh.cnrs.fr/infevers/) [30]. As a final verification of the sequences, we translated the mutant mRNA sequences using an automated translation tool (https://web.expasy.org/translate/) to ensure they matched the reported amino acids and positions from individual publications. Overall, this study did not require consent or approval from an Institutional Review Board as all mRNA and protein sequences were obtained or derived from publicly available sources: NCBI, publications, and the Infevers database. Our analysis focused solely on the mutations reported and their impacts on the structure and binding affinity of the resulting IL-1Ra protein.

**Table 1.**
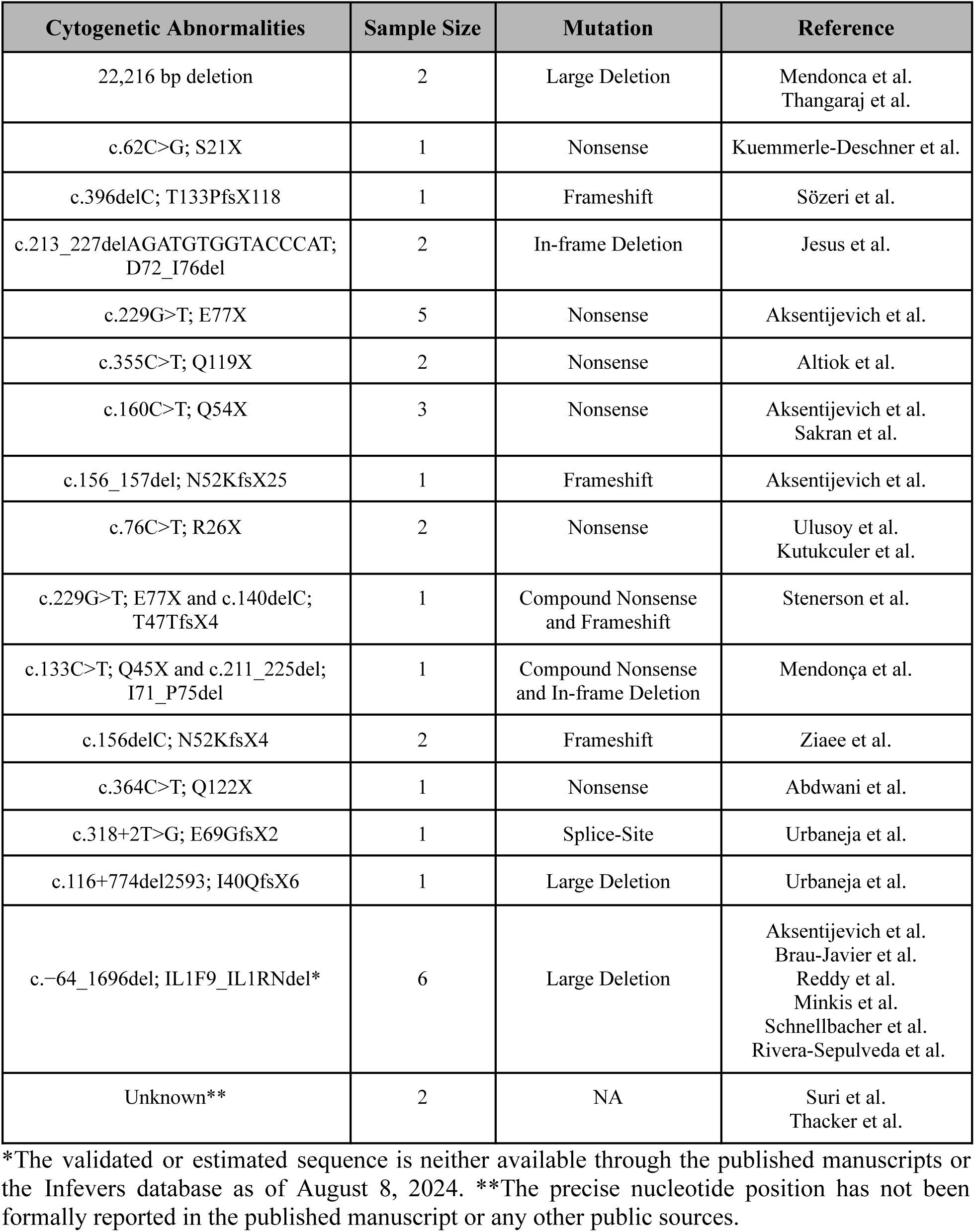
Cytogenetic abnormalities reported in DIRA patients (*n* = 34).

However, one limitation that occurred during sequence acquisition was that the large deletion variant of a 175-kb deletion was unable to be formulated. This was due to the complexity of the mutation, where it involved 6 different IL-1 related genes, as well as the lack of data availability and validation of the precise mRNA sequences from publications or Infevers database. Despite this limitation, this study was able to estimate the protein sequences of 15 variants.

### 2.2. 3D Structure Acquisition

Following acquisition of protein sequences, we followed the validated methodologies reported by Jesus et al. and Altiok et al. to predict alterations in secondary structure of the WT IL-1Ra protein *in silico* [13–14]. Both studies used the resolved protein structure of IL-1Ra in its binding mode to IL-1R1 from Schreuder et al. found with X-Ray crystallography [31]. Given the complexity of the mutations reported for DIRA as well as the fact that mutagenesis can only be evaluated for missense variants with current technological innovations, these studies naturally only highlighted the impacted regions of the protein, describing impacted β-strands, α-helices, and amino acids necessary for binding. In fact, these two studies did not delve further into detailed alterations in protein stability (i.e. free energy changes) or structural damages (e.g. bond breakages, etc). This study used this same protocol for 15 variants, 2 of which were completed by these studies, and will help validate the workflow of this study. It is important to note that the experimental model is missing the signal peptide cleavage as well as 5 amino acids following this sequence, but does not impact the structural analyses of this study or of the prior.

### 2.3. Predicting Structural Alterations

Within the WT IL-1Ra structure, the known structure is composed of 12 β-strands and 3-10 α-helices [32]. β1, β4, β5, β8, β9 and β12 are particularly important for the formation of the protein as they are critical to forming its six-stranded β-barrel [32]. From the PDB, the experimental model by Schreuder et al. reported 4 α-helices [31]. In order to streamline the process of identifying the secondary structures and impacted regions from individual sequences, we aligned the sequences and annotated the partial amino acid sequences to their corresponding secondary structure. In Supplementary Material 1, we have provided these annotated structures derived from the Schreuder et al. model.

After annotating all 15 sequences, we recorded the specific secondary structures that were retained from the experimental model to the mutant. We only considered the individual structures to be retained if all WT amino acids were present. Structures that contained partially correct sequences and additional novel amino acids were not considered a part of the count of original structures. Overall, from the 15 genetic variants, we have counted the β-strands and α-helices retained from the original model.

### 2.4. Predicting Alterations in Binding Sites

From a prior study by Evans et al., authors identified through site-directed mutagenesis that W16, Q20, Y34, Q36, and Y147 on IL-1Ra are essential for its binding affinity to IL-1R1, and later validated by the Schreuder et al. model [31, 33]. Evans et al. reported that upon binding of IL-1Ra, the free energy change of the entire structure was 52 kJ/mol, and found that 4 of the 5 sites (W16, Q20, Y34, and Y147) accounted for at least 18.3 kJ/mol (35.2%) [33].

At the binding interface of IL-1Ra, W16 forms hydrophobic interactions at domain 2 of the receptor, important for orientation and structural integrity of the cytokine. Q20 forms two hydrogen bonds with the main chain of the receptor on domain 2, critical for stabilization of the binding conformation. At the receptor side chain, Y34 similarly forms a hydrogen bond. Similarly, on domains 1 and 2 of IL-1R1, Q36 forms 3 hydrogen bonds likely for stabilization.

Lastly, Y147 does not directly interact with domain 2 of the receptor but uniquely with 2 water molecules to form a hydrogen bond with IL-1R1.

Overall, this study annotated the binding sites present in the WT IL-1Ra and its variants from the aligned sequences. This information has also been included in Supplementary Material 1. We have comprehensively counted the binding sites present in the sequences among the variants.

### 2.5. Validation of Structural Analyses

To further validate our findings of impacts on secondary structure and binding affinity, evidence from *in vitro* studies are necessary. However, considering the challenges of obtaining the individual patient data or performing a broad experimental protocol, it is not feasible to conduct these experiments and beyond the scope of this study. Yet, prior studies have already performed experiments that can help further validate the conclusions presented by this study. Within this work, we delve into the individual variants in the context of the *IL1RN* mRNA and subsequent protein expression from quantitative polymerase chain reaction (qPCR) and Western blotting experiments, respectively. If neither experiments were reported within studies, or any applicable experiments, we have noted this as well.

## 3. Conclusions

Within this study, we have conducted a structural analysis of impacted secondary structures and critical binding sites of IL-1Ra with IL-1R1 from the novel mutations reported in prior literature. In Fig. 1, we have provided counts of secondary structures, namely the α-helices and β-sheets in all structures. Likewise, within Fig. 2, we have provided counts of whether specific binding sites were still present in the variant structures for the 5 sites discussed previously. We also discuss relevant *in vitro* experiments performed from prior studies to support the findings of this work.

**Figure 1.**
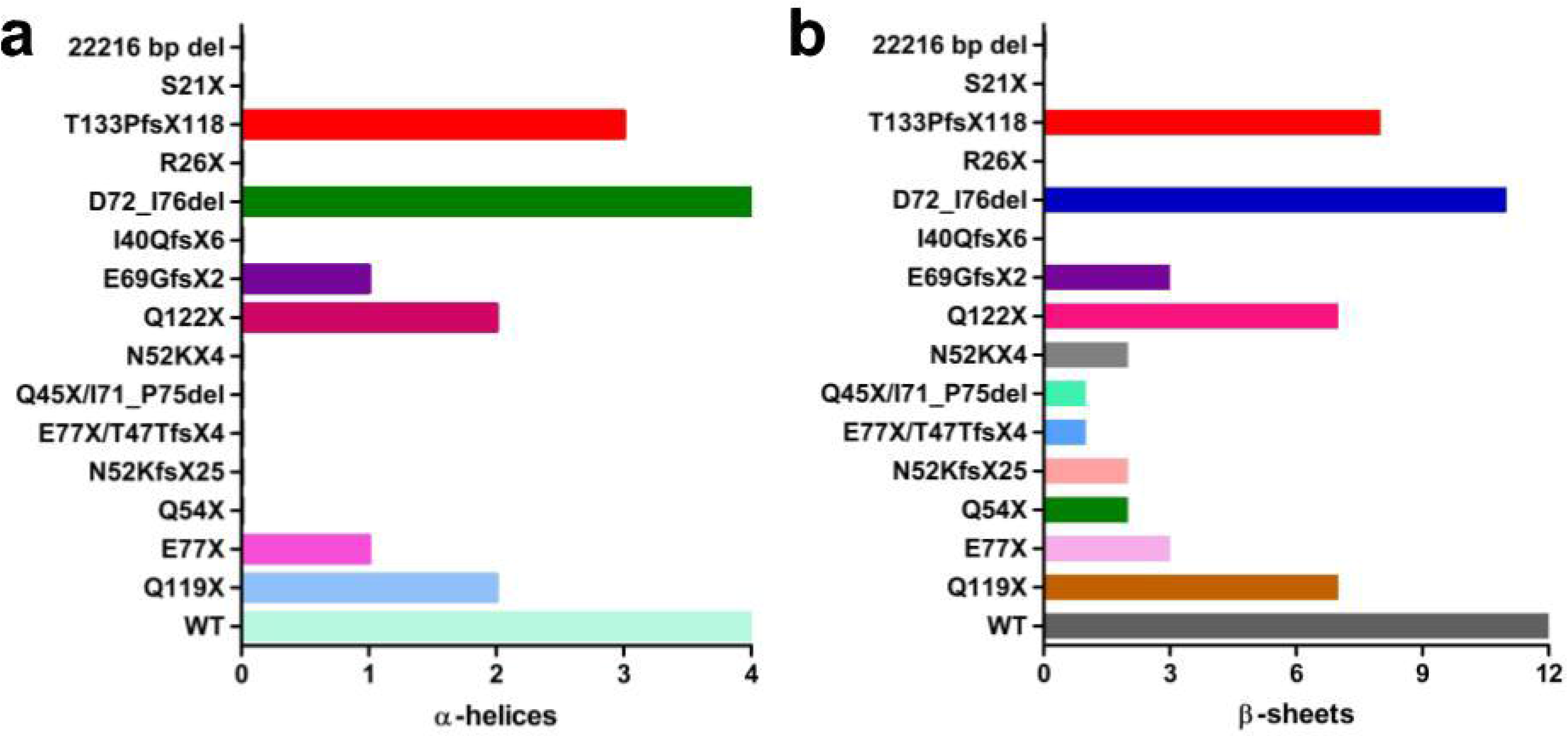

**Figure 2.**
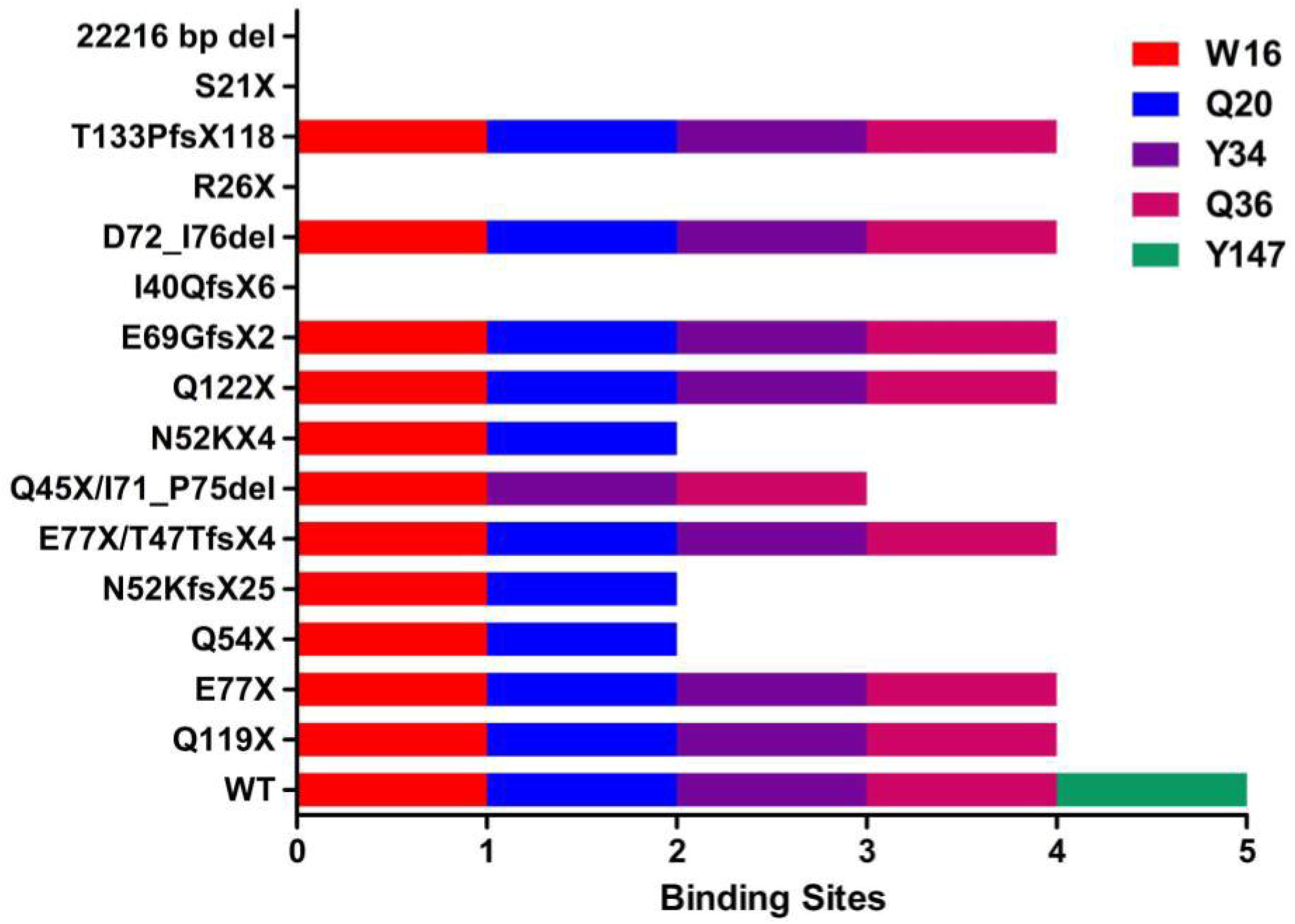

### 3.1. Nonsense Variants

Of the DIRA patients in literature, 6 unique nonsense variants have been reported up to date. The first variants were reported by Aksentijevich et al. including Q54X and E77X in 2009 [8]. From our prediction model, the Q54X variant would retain β1- and β2-strands and no α-helices, while E77X would have β1-, β2-, β3-strands and 1 α-helix. In terms of binding affinity, most sites were missing on Q54X, present with only W16 and Q20, while E77X was only missing Y147. Given that both variants were missing most critical secondary structures, and a significant number of binding sites, it would be expected that there would be no expression of the resulting proteins or at most of a non-functional truncated protein. From the clinical trial and the individual case report, unfortunately there were no experiments performed to evaluate the expression of the mRNA or protein expression of the Q54X variant [8, 15]. However, E77X was comprehensively evaluated in the clinical trial alone [8]. From qPCR, the heterozygote E77X variants showed diminished expression of *IL1RN* mRNA (*n* = 2), and even less in the homozygote (*n* = 1). In western blotting, a band corresponding to IL-1Ra was present but diminished in the heterozygotes, and completely missing in the homozygote.

Next, Ulusoy et al. reported a R26X variant in 2015 [16]. From the *in silico* model, it was predicted that no β-strands or α-helices would be retained from the WT, and therefore no binding sites would be present. Similarly, Kuemmerle-Deschner et al. reported a S21X variant in 2020, and had the same outcome of prediction [11]. The reasoning for these results is that the nonsense mutations occur in the signal peptide cleavage not present in the Schreuder et al. experimental model. Although the R26X had no *in vitro* experiments, S21X did have supporting data. From the S21X variant, there was induction of *IL1RN* mRNA expression, but no expression from the resulting IL-1Ra protein.

Lastly, the remaining two variants reported were by Abdwani et al. in 2021 and Altiok et al. in 2018 for a Q122X and Q119X variant, respectively [14, 21]. The prediction model displayed that the Q122X variant would retain β1-, β2-, β3-, β4-, β5-, β6-, and β7-strands with the first two α-helices, and therefore only missing the Y147 binding site. Unfortunately, this variant did not have any further experimental data presented. Conversely, the Q119X was predicted to retain the same secondary structures and binding sites as the Q122X variant. The potential explanation for these sequences being similar is that Q122X variant refers to different protein isoform that differs only in the segment prior to the signal peptide cleavage but retains the same amino acid sequences from the Schreuder et al. experimental model, and was the primary reason both structures were able to be predicted. Furthermore, our findings for the Q119X variant were validated by Altiok et al. that stated that the “β8-, β9- and β12-strands are missing” from the β-barrel of IL-1Ra [14].

### 3.2. Large Deletion and Splice-Site Variants

From the known DIRA patients, we predicted the alterations in structure and binding affinity of 2 unique large deletions and 1 splice-site variant. The first large deletion variant was first reported by Mendonca et al. in 2017 for a 22,216 bp deletion [9]. The authors reported that the mutation spanned the promoter region and the first 2 coding exons of the *IL1RN* gene, with no *in vitro* protein expression testing performed in the study. From our prediction of this variant, there would be no original β-strands, α-helices, or binding sites retained given the promoter sequence has been removed.

The other splice-site and large deletion variants, specifically E69GfsX2 and I40QfsX6, have been recently reported by Urbanaja et al. in 2024 [22]. For the E69GfsX2, the variant was predicted to retain the β1-, β2-, β3-strands and the first 1 α-helix, only missing the Y147 binding site total. Likewise, for the I40QfsX6 variant, no β-strands, α-helices, or binding sites are retained. From the study, the authors indirectly tested for protein expression, measuring *IL1RN* mRNA levels and gene expression from RNA-seq for both variant types. Both variants were found to have decreased levels of mRNA expression compared to the WT.

### 3.3 Frameshift Variants

There have been a total of 3 frameshift variants that have been reported for DIRA to date. The first variant was N52KfsX25, reported by Aksentijevich et al. in 2009 [8]. From the prediction model, the variant was expected to retain β1- and β2-strand but no α-helices, and W16 and Q20. Similarly to the 2 other variants reported from this clinical trial, analysis of *IL1RN* mRNA expression revealed diminished expression in the heterozygotes and even less in the homozygote. Likewise, western blotting revealed that the band of the heterozygotes were present but still diminished, and the homozygote being completely absent.

The other two variants reported, T133PfsX118 and N52KfsX4, were identified by Sözeri et al. and Ziaee et al. in 2018 and 2020, respectively [12, 20]. Unfortunately, no experimental studies were conducted in either of these case reports. From the T133PfsX118 variant, it is predicted that the structure would be missing β8-, β9-, β10-, β11-, and β12-strands and the last α-helix, ultimately retaining all major binding sites except Y147. For the N52KfsX4 variant, the structure is expected to only retain β1- and β2-strands, containing W16 and Q20.

### 3.4. In-frame Deletions & Compound Variants

Within current literature, there has been 1 variant with an in-frame deletion and 2 others reported for compound mutations. Jesus et al. first reported a D72_I76del variant in 2011, comprehensively reporting the expression of the resulting *IL1RN* mRNA and subsequent IL-1Ra [13]. From the prediction model, the deletion would impact most of the β4-strand in the WT structure. Like the data presented for other variants, mRNA expression was diminished in heterozygous parents, while the homozygotes themselves were even less. Interestingly, however, the variant was expressed in lower levels than the heterozygous parents and anakinra itself (positive control) during western blotting. However, from further tests, no binding was found between the mutant IL-1Ra and its IL-1R1. Additionally, our prediction analysis was also validated by the findings of this study, where the authors stated the “missing amino acids are in strand β4”.

Furthermore, the two compound variants were reported by Stenerson et al. in 2011 and Mendonca et al. in 2020 [18–19]. Stenerson et al. evaluated a E77X and unique T47TfsX4 variant, where only the β1-strand was retained and no α-helices, along with 2 binding sites (W16 and Q20). The case report for this patient did predict that the protein would be truncated to be ⅓ the size of the WT (177 amino acids), which was close to those predicted from this study of 49 amino acids. However, no other *in vitro* studies experiments were performed for this variant. Lastly, Mendonca et al. reported a variant containing a Q45X and I71_P75del mutation. From the structural analysis of this study, it retained the same number of original β-strands and α-helices to the other compound variant, but had only kept the W16 binding site.

## 4. Discussion

This brief communication detailed how the known mutations impacted critical binding sites of variants and secondary structures of IL-1Ra, causing DIRA disease. Among all variants, 80% of structures were predicted to retain W16, 73.3% would retain Q20, 60% would have Y34, 60% would retain Q36, and 0% would have Y147. From this data alone, it is expected that there is a reduction in binding affinity of IL-1Ra and IL-1R1, resulting in unopposed pro-inflammatory signaling. With the secondary structures among the variants, 7 variants are predicted to only retain the first 3 β-strands, and 4 are expected to retain more than 6 β-strands. In context of the β-barrel of WT IL-1Ra, all variants appear to have impacted β-strands, but Q119X, D72_I76del, Q122X, and T133PfsX118 all retain more than half of the necessary original structures. Therefore, these findings reveal critical structural impacts in secondary structure, likely to have an impact in the function of the resulting IL-1Ra proteins.

Among the nonsense variants, 3 variants had supporting evidence from *in vitro* data. The E77X and S21X variants were shown to have diminished expression of the *IL1RN* mRNA, and subsequently no expression of the IL-1Ra protein. The findings of the Q119X variant were replicated in this study from the Altiok et al. case report. Nonsense variants remain the prevalent variant type among others, and the findings from this study show that major truncations in protein structure will impact binding affinity and secondary structure, leading to no resulting expression. For the large deletion and splice-site variants, these mutations appear to have a significant impact on the resulting protein translation. Although not evaluated in this study directly, the 175-kb deletion variant was found to have no protein expression from the Aksentijevich et al. led clinical trial [8]. This is a similar trend observed with the other variants reported, specifically I40QfsX6 and the 22,216 bp deletion, that had significantly impacted structures, but E69GfsX2 still remains at odds with this trend.

For the frameshift variants, only 1 was directly evaluated *in vitro*, and was similarly found to have no protein expression. Lastly, and most interestingly, for the in-frame deletions, the variant by Jesus et al. was found to have expression of the IL-1Ra. This variant retained the most secondary structures and binding sites when compared to the other variants, as only the β4-strand was impacted. This variant was the only known mutation reported to still have expression of the truncated IL-1Ra protein compared to all others that yielded no expression. However, further studies are still required to evaluate the resulting functional alterations from changes in structure and binding affinity.

We believe that the broad structural analyses performed in this study provides valuable insights into quantifying the impacts of all variants on the structure and binding of IL-1Ra. To the best of our knowledge, there are no prior studies that have reported these alterations. The findings of this study may be informative to clinicians diagnosing, treating, and maintaining DIRA, especially when the variant has been thoroughly reported in prior literature. Although there is still much information unknown of the function and stability of resulting IL-1Ra proteins, the data reported from this study remain valuable for understanding this rare disease and its pathogenic impacts.

However, there are some limitations of this brief communication that must be noted. One limitation with this study is that not all variants had *in vitro* data from experimental studies. Although performing these tests to understand the resulting function of IL-1Ra from the predicted impacts are important, these evaluations are beyond the scope of this brief communication. Our workflow used in this study was rooted from the methodologies reported by Jesus et al. and Altiok et al., allowing for us to confidently predict impacted secondary structures as well as binding sites. From our results, we accurately validated the findings from both of these studies for the structural impacts. Therefore, while supporting *in vitro* data is beneficial, it does not fundamentally alter the workflow used in this study. We recommend a more comprehensive experimental analysis for studies in the future.

Another shortcoming of this brief communication was that we did not comprehensively record all predicted alterations in binding sites among the variants. In order to streamline the process of evaluating binding affinity and to maintain uniformity in analysis of variants, this study focused on the 5 critical sites previously reported only. A broad analysis of all molecular interactions between IL-1Ra and IL-1R1 for known variants is required for future studies, and especially changes in free energy.

## 5. Conclusion

In this brief report, we used the methodology first used by Jesus et al. and Altiok et al. to evaluate impacts from mutations on the resulting structure and binding affinity of IL-1Ra with its receptor, IL-1R1, for 15 known variants reported for DIRA disease. We found that there were decreased binding affinities as 5 critical binding sites (W16, Q20, Y34, Q36, and Y147). Secondly, structural analyses revealed that significant secondary structures were impacted, likely to alter the stability and function of the resulting IL-1Ra protein. With the rarity of these variants alone and the varying levels of data reporting, we also discussed the *in vitro* studies previously performed for 7 individual variants. Although further experimental studies are required for studying this challenging and life-threatening disease, the *in silico* findings from this study may be informative to efforts to diagnose, treat, and maintain this disease as well as understanding its pathogenic mechanisms.

## Data Availability

All supporting data and analysis is available in Supplementary Materials 1.

## Acknowledgements

Not applicable

## Author Contributions

J.P. conducted all aspects of this study.

## Data Availability

The data supporting the results of this study are provided in Supplementary Material 1.

## Abbreviations

IL-1Ra: Interleukin-1 Receptor Antagonist
DIRA: Deficiency of Interleukin-1 Receptor Antagonist
NCBI: National Center for Biotechnology Information
IL-1R1: IL-1 Receptor Type 1
PDB: Protein Data Bank
WT: Wild Type

