## Supplementary Material 1 for "Alterations in secondary structure and binding affinity from mutations in the interleukin-1 receptor antagonist": Supplementary Material 1.pdf

**Joshua Pillai**[1, 2*]

[1]School of Biological Sciences, University of California, San Diego, 9301 S Scholars Dr, La Jolla, 92093, CA, USA.

[2]Department of Neurosciences, University of California, San Diego, School of Medicine, 9500 Gilman Dr, La Jolla, 92093, CA, USA.

*To whom correspondence should be addressed: Joshua Pillai

**This PDF file includes:**

Supplementary Note 1-16

**Supplementary Note 1 - Secondary Structures and Binding Sites from WT**

β1 - QAFRIWD

β2 - TFYLR

β3 - QLVAG

β4 - IDVVPIE

β5 - ALFLG

β6 - CLSCVKS

β7 - ETRLQLE

β8 - AFIRSDS

β9 - TTSFES

β10 - FLC

β11 - SLT

β12 - FYFQE

α1 - GPNVNL

α2 - ITD

α3 - KQDKRF

α4 - PDE

Validation of Protein Sequence

https://www.rcsb.org/structure/1IRA

**NOTE:** Residues 1-25 are part of signal peptide cleavage and "RPSGRK" is missing from the experimental model. The model will begin at the following amino acids "SSKM".

**Supplementary Note 2 - Altiok et al.**

```
                    10         20         30         40
WT          MEICRGLRSH LITLLLFLFH SETICRPSGR KSSKMQAFRI
Q119X       MEICRGLRSH LITLLLFLFH SETICRPSGR KSSKMQAFRI
            ****************************************

                    50         60         70         80
WT          WDVNQKTFYL RNNQLVAGYL QGPNVNLEEK IDVVPIEPHA
Q119X       WDVNQKTFYL RNNQLVAGYL QGPNVNLEEK IDVVPIEPHA
            ****************************************

                    90        100        110        120
WT          LFLGIHGGKM CLSCVKSGDE TRLQLEAVNI TDLSENRKQD
Q119X       LFLGIHGGKM CLSCVKSGDE TRLQLEAVNI TDLSENRK
            ****************************************

                   130        140        150        160
WT          KRFAFIRSDS GPTTSFESAA CPGWFLCTAM EADQPVSLTN
            ****************************************

                   170
WT          MPDEGVMVTK FYFQEDE
            *****************
```

<u>Notes</u>

β8, β9, β10, β11, and β12 are missing. For α-helices, the third and fourth are missing. With binding sites, only Y147 is missing.

<u>Validation of Protein Sequence</u>

Publication reporting verifies predicted missing secondary structure units.

<u>Reference</u>

Altiok, E. *et al.* A novel mutation in the interleukin-1 receptor antagonist associated with intrauterine disease onset. *Clinical Immunology* **145**, 77–81 (2012).

**Supplementary Note 3 - Aksentijevich et al.**

```
                  10         20         30         40
WT         MEICRGLRSH LITLLLFLFH SETICRPSGR KSSKMQAFRI
E77X       MEICRGLRSH LITLLLFLFH SETICRPSGR KSSKMQAFRI
           ****************************************

                  50         60         70         80
WT         WDVNQKTFYL RNNQLVAGYL QGPNVNLEEK IDVVPIEPHA
E77X       WDVNQKTFYL RNNQLVAGYL QGPNVNLEEK IDVVPI
           ****************************************

                  90        100        110        120
WT         LFLGIHGGKM CLSCVKSGDE TRLQLEAVNI TDLSENRKQD
           ****************************************

                 130        140        150        160
WT         KRFAFIRSDS GPTTSFESAA CPGWFLCTAM EADQPVSLTN
           ****************************************

                 170
WT         MPDEGVMVTK FYFQEDE
           *****************
```

<u>Notes</u>

β4 is partially completed. β5-12 are missing entirely. α-helices 2-4 are missing. For interactions, the only missing binding site is Y147.

<u>Validation of Protein Sequence</u>

Supplementary material of the publication displays the reported MSA of protein sequence.

<u>Reference</u>

Aksentijevich, I. *et al.* An Autoinflammatory Disease with Deficiency of the Interleukin-1–Receptor Antagonist. *New England Journal of Medicine* **360**, 2426–2437 (2009).

**Supplementary Note 4 - Aksentijevich et al.**

```
                   10         20         30         40
WT        MEICRGLRSH LITLLLFLFH SETICRPSGR KSSKMQAFRI
Q54X      MEICRGLRSH LITLLLFLFH SETICRPSGR KSSKMQAFRI
          ****************************************

                   50         60         70         80
WT        WDVNQKTFYL RNNQLVAGYL QGPNVNLEEK IDVVPIEPHA
Q54X      WDVNQKTFYL RNN
          ****************************************

                   90        100        110        120
WT        LFLGIHGGKM CLSCVKSGDE TRLQLEAVNI TDLSENRKQD
          ****************************************

                  130        140        150        160
WT        KRFAFIRSDS GPTTSFESAA CPGWFLCTAM EADQPVSLTN
          ****************************************

                  170
WT        MPDEGVMVTK FYFQEDE
          *****************
```

Notes

β3-12 are missing. α-helices 1-4 are missing. For interactions, the missing binding sites are Y34, Q36, and Y147.

Validation of Protein Sequence

Supplementary material of the publication displays the reported MSA of protein sequence.

**Supplementary Note 4 - Aksentijevich et al.**

```
                                10         20         30         40
WT              MEICRGLRSH LITLLLFLFH SETICRPSGR KSSKMQAFRI
N52KfsX25       MEICRGLRSH LITLLLFLFH SETICRPSGR KSSKMQAFRI
                ******************************************

                                50         60         70         80
WT              WDVNQKTFYL RNNQLVAGYL QGPNVNLEEK IDVVPIEPHA
N52KfsX25       WDVNQKTFYL RKPTSCWILA RTKCQFRRKD RCGTH
                ******************************************

                                90        100        110        120
WT              LFLGIHGGKM CLSCVKSGDE TRLQLEAVNI TDLSENRKQD
                ******************************************

                               130        140        150        160
WT              KRFAFIRSDS GPTTSFESAA CPGWFLCTAM EADQPVSLTN
                ******************************************

                               170
WT              MPDEGVMVTK FYFQEDE
                ******************
```

Notes

β3-12 are missing. α-helices 1-4 are missing. For interactions, the missing binding sites are Y34, Q36, and Y147.

Validation of Protein Sequence

Supplementary material of the publication displays the reported MSA of protein sequence.

**Supplementary Note 6 - Stenerson et al.**

```
                               10         20         30         40
WT                     MEICRGLRSH LITLLLFLFH SETICRPSGR KSSKMQAFRI
E77X/T47TfsX4          MEICRGLRSH LITLLLFLFH SETICRPSGR KSSKMQAFRI
                       ****************************************

                               50         60         70         80
WT                     WDVNQKTFYL RNNQLVAGYL QGPNVNLEEK IDVVPIEPHA
E77X/T47TfsX4          WDVNQKTSI
                       ****************************************

                               90        100        110        120
WT                     LFLGIHGGKM CLSCVKSGDE TRLQLEAVNI TDLSENRKQD
                       ****************************************

                              130        140        150        160
WT                     KRFAFIRSDS GPTTSFESAA CPGWFLCTAM EADQPVSLTN
                       ****************************************

                              170
WT                     MPDEGVMVTK FYFQEDE
                       *****************
```

Notes

β2 is partial. β3-12 is missing. α-helices 1-4 are missing. For interactions, the missing binding sites are Y34, Q36, and Y147.

Validation of Protein Sequence

Publication reports c.140delC; p.T47TfsX4 but Infevers reports c.141del; p.Phe48Serfs*3. Both appear to be correct and the sequence obtained from this study supports it.

**Supplementary Note 7 - Mendonca et al.**

```
                             10         20         30         40
WT                    MEICRGLRSH LITLLLFLFH SETICRPSGR KSSKMQAFRI
Q45X/I71_P75del       MEICRGLRSH LITLLLFLFH SETICRPSGR KSSKMQAFRI
                      ***************************************

                             50         60         70         80
WT                    WDVNQKTFYL RNNQLVAGYL QGPNVNLEEK IDVVPIEPHA
Q45X/I71_P75del       WDVN
                      ***************************************

                             90        100        110        120
WT                    LFLGIHGGKM CLSCVKSGDE TRLQLEAVNI TDLSENRKQD
                      ***************************************

                            130        140        150        160
WT                    KRFAFIRSDS GPTTSFESAA CPGWFLCTAM EADQPVSLTN
                      ***************************************

                            170
WT                    MPDEGVMVTK FYFQEDE
                      *****************
```

Notes

β2-12 are missing. α-helices 1-4 are missing. For interactions, the missing binding sites are Q20, Y34, Q36, and Y147.

Validation of Protein Sequence

Supplementary material and Sanger sequencing supports protein sequence reported.

**Supplementary Note 8 - Ziaee et al.**

```
                            10         20         30         40
WT              MEICRGLRSH LITLLLFLFH SETICRPSGR KSSKMQAFRI
N52KfsX4        MEICRGLRSH LITLLLFLFH SETICRPSGR KSSKMQAFRI
                ****************************************

                            50         60         70         80
WT              WDVNQKTFYL RNNQLVAGYL QGPNVNLEEK IDVVPIEPHA
N52KfsX4        WDVNQKTFYL RKTN
                ****************************************

                            90        100        110        120
WT              LFLGIHGGKM CLSCVKSGDE TRLQLEAVNI TDLSENRKQD
                ****************************************

                           130        140        150        160
WT              KRFAFIRSDS GPTTSFESAA CPGWFLCTAM EADQPVSLTN
                ****************************************

                           170
WT              MPDEGVMVTK FYFQEDE
                *****************
```

<u>Notes</u>

β3-12 are missing. α-helices 1-4 are missing. For interactions, the missing binding sites are Y34, Q36, and Y147.

<u>Validation of Protein Sequence</u>

Paper reports c.54delC;p.Asn18Lysfs*4 but Infevers reports c.156delC;p.Asn52Lysfs*4. There appears to be an error in the reporting of the publication. The mRNA sequence also does not align from the publication and there is no cytosine at position 54 after the start codon. However, it is correctly aligned in the Infevers reporting. This study will use the reported mutation from Infevers. The author will contact the corresponding authors of this study to verify the mutation and sequence.

<u>Reference</u>

Ziaee, V. *et al.* Homozygous IL1RN Mutation in Siblings with Deficiency of Interleukin-1 Receptor Antagonist (DIRA). *Journal of Clinical Immunology* **40**, 637–642 (2020)

**Supplementary Note 9 - Abdwani et al.**

```
                  10         20         30         40
WT         MALADLYEEG GGGGGEGEDN ADSKETICRP SGRKSSKMQA
Q122X      MALADLYEEG GGGGGEGEDN ADSKETICRP SGRKSSKMQA
           ****************************************

                  50         60         70         80
WT         FRIWDVNQKT FYLRNNQLVA GYLQGPNVNL EEKIDVVPIE
Q122X      FRIWDVNQKT FYLRNNQLVA GYLQGPNVNL EEKIDVVPIE
           ****************************************

                  90        100        110        120
WT         PHALFLGIHG GKMCLSCVKS GDETRLQLEA VNITDLSENR
Q122X      PHALFLGIHG GKMCLSCVKS GDETRLQLEA VNITDLSENR
           ****************************************

                 130        140        150        160
WT         KQDKRFAFIR SDSGPTTSFE SAACPGWFLC TAMEADQPVS
Q122X      K
           ****************************************

                 170
WT         LTNMPDEGVM VTKFYFQEDE
           *********************
```

Notes

β8-12 are missing. α-helices 4 is missing. For interactions, the missing binding sites are Y34, Q36, and Y147.

Validation of Protein Sequence

None possible from public information. Although the mRNA sequence varies, after alternative splicing the protein sequence becomes aligned after the first 34 residues.

**Supplementary Note 10 - Urbaneja et al.**

```
                    10         20         30         40
WT          MEICRGLRSH LITLLLFLFH SETICRPSGR KSSKMQAFRI
E69GfsX2    MEICRGLRSH LITLLLFLFH SETICRPSGR KSSKMQAFRI
            ****************************************

                    50         60         70         80
WT          WDVNQKTFYL RNNQLVAGYL QGPNVNLEEK IDVVPIEPHA
E69GfsX2    WDVNQKTFYL RNNQLVAGYL QGPNVNLEGS
            ****************************************

                    90        100        110        120
WT          LFLGIHGGKM CLSCVKSGDE TRLQLEAVNI TDLSENRKQD
            ****************************************

                   130        140        150        160
WT          KRFAFIRSDS GPTTSFESAA CPGWFLCTAM EADQPVSLTN
            ****************************************

                    170
WT          MPDEGVMVTK FYFQEDE
            *****************
```

Notes

β4-12 are missing. α-helices 2-4 are missing. For interactions, the missing binding site is Y147.

Validation of Protein Sequence

Supplementary material of the publication displays the reported protein sequence.

**Supplementary Note 11 - Urbaneja et al (Ile40Glnfs*6)**

```
                    10         20         30         40
WT            MEICRGLRSH LITLLLFLFH SETICRPSGR KSSKMQAFRI
I40QfsX6      MEICRGLRSH LITLLLFLFH SETICRPSGR KSSKMQAFRQ
              ****************************************

                    50         60         70         80
WT            WDVNQKTFYL RNNQLVAGYL QGPNVNLEEK IDVVPIEPHA
I40QfsX6      LTSLT
              ****************************************

                    90        100        110        120
WT            LFLGIHGGKM CLSCVKSGDE TRLQLEAVNI TDLSENRKQD
              ****************************************

                   130        140        150        160
WT            KRFAFIRSDS GPTTSFESAA CPGWFLCTAM EADQPVSLTN
              ****************************************

                    170
WT            MPDEGVMVTK FYFQEDE
              *****************
```

Notes

β1 is partial. β2-12 are completely missing. α-helices 1-4 are missing. For interactions, the missing binding sites are W16, Q20, Y34, Q36, and Y147.

Validation of Protein Sequence

Supplementary material of the publication displays the reported protein sequence.

**Supplementary Note 12 - Jesus et al.**

```
                           10         20         30         40
WT                  MEICRGLRSH LITLLLFLFH SETICRPSGR KSSKMQAFRI
D72_I76del          MEICRGLRSH LITLLLFLFH SETICRPSGR KSSKMQAFRI
                    *******************************************

                           50         60         70         80
WT                  WDVNQKTFYL RNNQLVAGYL QGPNVNLEEK IDVVPIEPHA
D72_I76del          WDVNQKTFYL RNNQLVAGYL QGPNVNLEEK IEPHALFLGI
                    *******************************************

                           90        100        110        120
WT                  LFLGIHGGKM CLSCVKSGDE TRLQLEAVNI TDLSENRKQD
D72_I76del          HGGKMCLSCV KSGDETRLQL EAVNITDLSE NRKQDKRFAF
                    *******************************************

                          130        140        150        160
WT                  KRFAFIRSDS GPTTSFESAA CPGWFLCTAM EADQPVSLTN
D72_I76del          IRSDSGPTTS FESAACPGWF LCTAMEADQP VSLTNMPDEG
                    *******************************************

                          170
WT                  MPDEGVMVTK FYFQEDE
D72_I76del          VMVTKFYFQE DE
                    *****************
```

Notes

β1-12 are completely missing. α-helices 1-4 are missing. For interactions, the missing binding sites are W16, Q20, Y34, Q36, and Y147.

Validation of Protein Sequence

Manuscript reporting verifies predicted missing secondary structure units.

**Supplementary Note 13 - Ulusoy et al.**

```
                  10         20         30         40
WT        MEICRGLRSH LITLLLFLFH SETICRPSGR KSSKMQAFRI
R26X      MEICRGLRSH LITLLLFLFH SETICRPSG
          *******************************************

                  50         60         70         80
WT        WDVNQKTFYL RNNQLVAGYL QGPNVNLEEK IDVVPIEPHA
          *******************************************

                  90        100        110        120
WT        LFLGIHGGKM CLSCVKSGDE TRLQLEAVNI TDLSENRKQD
          *******************************************

                 130        140        150        160
WT        KRFAFIRSDS GPTTSFESAA CPGWFLCTAM EADQPVSLTN
          *******************************************

                 170
WT        MPDEGVMVTK FYFQEDE
          ******************
```

Notes

β1-12 are completely missing. α-helices 1-4 are missing. For interactions, the missing binding sites are W16, Q20, Y34, Q36, and Y147.

Validation of Protein Sequence

The sequence is verified by the Infevers database. Limited information is provided of the cytogenetic abnormality in the publication.

**Supplementary Note 14 - Sozeri et al.**

```
                          10         20         30         40
WT              MEICRGLRSH LITLLLFLFH SETICRPSGR KSSKMQAFRI
T133PfsX118     MEICRGLRSH LITLLLFLFH SETICRPSGR KSSKMQAFRQ
                ****************************************

                          50         60         70         80
WT              WDVNQKTFYL RNNQLVAGYL QGPNVNLEEK IDVVPIEPHA
T133PfsX118     WDVNQKTFYL RNNQLVAGYL QGPNVNLEEK IDVVPIEPHA
                ****************************************

                          90        100        110        120
WT              LFLGIHGGKM CLSCVKSGDE TRLQLEAVNI TDLSENRKQD
T133PfsX118     LFLGIHGGKM CLSCVKSGDE TRLQLEAVNI TDLSENRKQD
                ****************************************

                         130        140        150        160
WT              KRFAFIRSDS GPTTSFESAA CPGWFLCTAM EADQPVSLTN
T133PfsX118     KRFAFIRSDS GPPPVLSLPP APVGSSAQRW KLTSPSASPI
                ****************************************

                         170        180        190        200
WT              MPDEGVMVTK FYFQEDE
T133PfsX118     CLTKASWSPN STSRRTSSTA QACLFPFLHG KDCRDCQSPC
                ****************************************

                         210        220        230        240
T133PfsX118     PRAPGYGGTE DQPLRGGPSE GVTTTWSQDS ASSSTDQPPC
                ****************************************


T133PfsX118     CLQNGLSNV
                *********
```

Notes

β8-12 are completely missing. α-helices 4 is missing. For interactions, the missing binding site is Y147.

Validation of Protein Sequence

From the verified mRNA sequence, we obtained the following protein sequence. Thr appears to be deleted and the sequence continues 117 amino acids. The paper mentions 118 amino acids downstream. Also, regarding the polymorphism, cytosine appears to be in the isoform variant 1 and not thymine. The author will contact the corresponding author of the study to inquire about the sequence.

**Supplementary Note 15 - Kuemmerle-Deschner et al.**

```
                 10         20         30         40
WT       MEICRGLRSH LITLLLFLFH SETICRPSGR KSSKMQAFRI
S21X     MEICRGLRSH LITLLLFLFH
         ****************************************

                 50         60         70         80
WT       WDVNQKTFYL RNNQLVAGYL QGPNVNLEEK IDVVPIEPHA
         ****************************************

                 90        100        110        120
WT       LFLGIHGGKM CLSCVKSGDE TRLQLEAVNI TDLSENRKQD
         ****************************************

                130        140        150        160
WT       KRFAFIRSDS GPTTSFESAA CPGWFLCTAM EADQPVSLTN
         ****************************************

                170
WT       MPDEGVMVTK FYFQEDE
         *****************
```

Notes

β1-12 are completely missing. α-helices 1-4 are missing. For interactions, the missing binding sites are W16, Q20, Y34, Q36, and Y147.

Validation of Protein Sequence

The sequence is verified by the Infevers database.

**Supplementary Note 16 - Mendonca et al (22,216 bp deletion)**

```
                         10         20         30         40
WT               MEICRGLRSH LITLLLFLFH SETICRPSGR KSSKMQAFRI
22,216 bp del    CQFRRKDRCG TH
                 *******************************************

                         50         60         70         80
WT               WDVNQKTFYL RNNQLVAGYL QGPNVNLEEK IDVVPIEPHA
                 *******************************************

                         90        100        110        120
WT               LFLGIHGGKM CLSCVKSGDE TRLQLEAVNI TDLSENRKQD
                 *******************************************

                        130        140        150        160
WT               KRFAFIRSDS GPTTSFESAA CPGWFLCTAM EADQPVSLTN
                 *******************************************

                        170
WT               MPDEGVMVTK FYFQEDE
                 ******************
```

Notes

β1-12 are completely missing. α-helices 1-4 are missing. For interactions, the missing binding sites are W16, Q20, Y34, Q36, and Y147.

Validation of Protein Sequence

Manually verified on NCBI. Using NC_000002.11, we modified the coordinates to 113,865,011 to 113,887,227 to identify the deleted region. Coding exon 1 is deleted completely and exon 2 almost completely except for "TGTCAATTTAGAAG". Coding exons 3 and 4 are still present. However, since the start codon is gone, the entire protein folding and sequence is gone and completely altered from the original structure. We have provided annotations of regions of the gene impacted in the data repository.
